# Potential adverse effects of an educational intervention: Development of a framework

**DOI:** 10.1101/2022.07.27.22278097

**Authors:** Matt Oxman, Faith Chelagat Chesire, Michael Mugisha, Ronald Ssenyonga, Benson Ngatia, Allen Nsangi, Simon Lewin, Jenny Moberg, Nelson Sewankambo, Margaret Kaseje, Monica Melby-Lervåg, Atle Fretheim, Andrew David Oxman, Sarah Rosenbaum

## Abstract

**Background:** Researchers often overlook potential adverse effects of educational and public health interventions (increases in adverse outcomes, or decreases in beneficial outcomes, attributed to the intervention). To help us identify potential adverse effects of an educational intervention intended to improve critical thinking about health choices, we developed a framework. We also did a preliminary prioritisation of outcomes in the framework for randomised trials of the intervention, and associated process evaluations.

**Methods:** Based on relevant evidence and theory, we developed an initial framework. For feedback on the initial framework, we sent a survey to 70 external experts. We conducted a thematic analysis of the qualitative survey data. After revising the framework based on the survey findings, we interviewed teachers in the context where we are evaluating the intervention, to help identify any effects still missing from the framework, and preliminarily prioritise potential outcomes for the evaluation.

**Results:** We received responses from 38 of the 70 external experts (54%), including researchers and others with a variety of expertise within health, education, and design. Overall, the responses were positive. However, they also included critical feedback that led to substantial revisions of the framework’s content and presentation. The revised framework has six categories of potential adverse effects: decision-making harms, psychological harms, equity harms, group and social harms, waste, and other harms. We interviewed three teachers, who did not suggest any missing outcomes. Based on the interview findings, we prioritised three outcomes for the evaluation of the intervention: work-related stress; wasted time or resources; and conflict, in particular between students and family.

**Discussion:** As far as we are aware, the framework presented in this article is the first tool of its kind in education research. The framework is a “living” tool, which can be improved upon, as well as adapted. We have used it to inform the development of interview and observation guides, and we are using it to inform the development of outcome measures. Important limitations of the framework include limits to its comprehensiveness, and the use of terminology with different meanings or interpretations depending on the context. Our approach to identifying and evaluating potential adverse effects of an educational intervention can have value to other researchers.

**Conclusion:** Rigorous evaluations of potential adverse effects of educational and public health interventions can be time and resource-intensive. However, that cost might be small compared to the cost of implementing harmful interventions.

## Background

### Adverse effects in education and public health

Educational and public health interventions warrant rigorous development and evaluation, especially prior to large-scale implementation—not only because the intervention might be ineffective and wasteful, but because it might cause adverse effects (increases in adverse outcomes or decreases in beneficial outcomes). However, researchers often overlook potential adverse effects of educational and public health interventions [1–4]. When they evaluate intended effects, this might reveal a lack of effect or paradoxical effect, but not other adverse effects (“side effects”) [1–4].

To help evaluate potential adverse effects of public health interventions, Lorenc and Oliver have developed a framework with categories and examples [3]. Building on said framework, Bonell et al. have proposed strategies for theorising how such effects might happen and creating “Dark logic” models [4]. While Zhao provides examples of adverse effects of educational interventions [1, 2], there does not appear to be a tool for education like the framework developed by Lorenc and Oliver for public health.

### Informed Health Choices project

As part of the Informed Health Choices (IHC) project (www.informedhealthchoices.org), we have developed and are evaluating an intervention to help secondary school students learn how to think critically about health information and choices. We are building on findings and experiences from developing and evaluating a corresponding intervention targeted at primary school children [5–7]. In .Box 1, we describe the aim, development and evaluation of the IHC secondary school intervention in more detail.

#### Box 1. Aim, development, and evaluation of the IHC secondary school intervention.

The IHC secondary school intervention is primarily aimed at improving students’ ability to apply a prioritised set of concepts [8]. The concepts come from the “Key Concepts for Informed Health Choices” framework [9], also known as the IHC Key Concepts framework. The ability to apply the IHC Key Concepts is part of “health literacy”, more specifically “critical health literacy” [10]. We developed the intervention in Kenya, Rwanda, and Uganda [11]. To inform the development, we first studied the contexts for teaching critical thinking about health using digital resources in those countries [12–14]. The final intervention has two components: training teachers and providing them with digital resources. After the training, teachers are intended to deliver 10 lessons to students. The lessons are introduced in the training and outlined in the resources. Each lesson focuses on one or more of the prioritised concepts. The intervention is both an educational and public health intervention in that we intend for it to improve educational outcomes, but also—in the long term, as part of a potential series of interventions—public health outcomes. We are evaluating the intervention in respective randomised trials in Kenya, Rwanda, and Uganda [15–17], and process evaluations associated with the trials.

In the trial of the IHC primary school intervention, participants did not report any adverse outcomes, although we asked the teachers to record and report any they experienced or observed [5, 18]. In the process evaluation associated with that trial, we did not observe any adverse outcomes, but some teachers reported an increase in stress due to the added workload, as well as teaching something new [6]. Furthermore, a majority of teachers and parents expressed concern that the intervention might cause conflict between themselves and the children, by causing children to challenge authority. There were reports of children challenging authority, but not of conflicts.

Like with the primary school intervention, we have employed an iterative, human-centred design approach to develop the secondary school intervention, involving cycles of prototyping, user-testing, and piloting. This approach is helpful for finding and addressing problems during the development stage that could potentially lead to adverse effects [7, 11]. Moreover, findings about potential adverse effects from the development and evaluation of the primary school intervention have informed the development of the secondary school intervention.

Given 1) the findings from the evaluation of the primary school intervention, 2) our approach to developing the secondary school intervention, and 3) use of what we have learned from the development and evaluation of the primary school intervention, we do not anticipate adverse effects of the secondary school intervention.

However, there were limitations to the evaluation of potential adverse effects of the primary school intervention. For example, we did not consult external experts about outcomes to measure or explore.

Despite our approach and using what we have learned, the secondary school intervention might have adverse effects that we have failed to identify or address, for example because of differences between schools that have participated in its development versus other schools. The bottom line: adverse effects are always possible.

### This study

To help prevent and evaluate potential adverse effects of any intervention, it is key to first identify potential adverse effects that are logical and likely to be important to participants or other stakeholders. Therefore, we developed a framework of potential adverse effects of the IHC secondary school intervention. We preliminarily prioritised outcomes in the framework to measure and explore in the trials and process evaluations. Fig 1 is an overview of the development and evaluation of the intervention, including this study.

**Fig 1.**
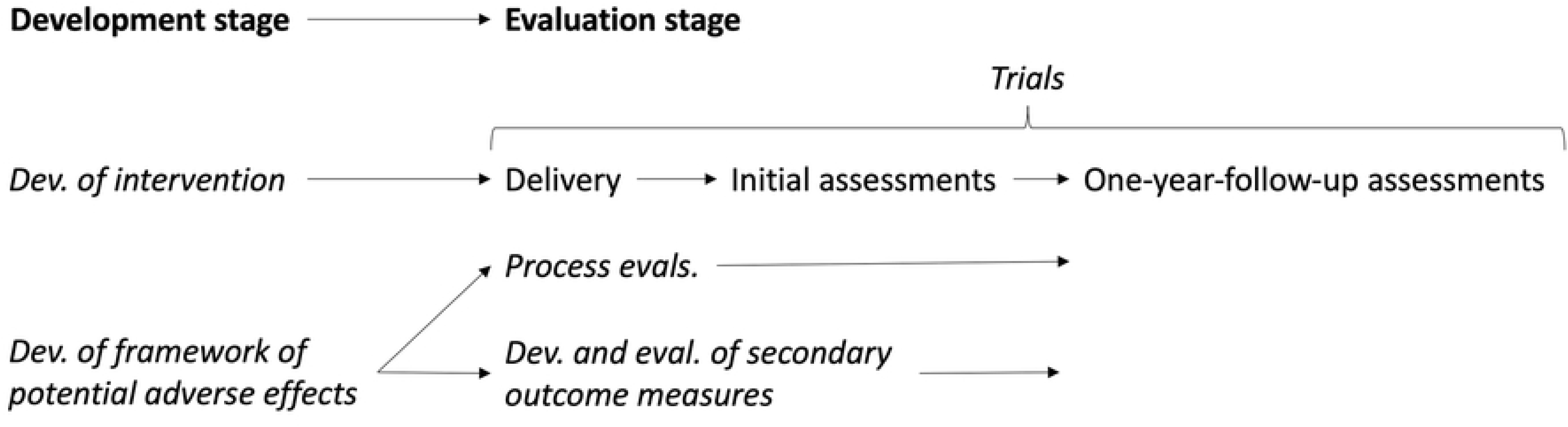
Development and evaluation of IHC secondary school intervention.

### Objectives

#### Primary

To identify potential adverse effects of the IHC secondary school intervention, by developing a framework of such effects

#### Secondary

To prioritise potential adverse outcomes included in the framework, for the evaluation of the intervention

## Methods

### Development of initial framework and criteria

We first developed an initial framework of potential adverse effects of the IHC secondary school intervention, including descriptions of their potential mechanisms and corresponding beneficial effects, as well as criteria for a sensible framework (S1 File).

We described the potential mechanisms to help identify potential adverse effects, as suggested by Bonell et al. [4], but also to help identify corresponding beneficial effects, and inform the development of the intervention towards preventing the potential adverse effects and achieving the corresponding beneficial effects. We based the first version of the framework on relevant evidence and theory, in particular: the framework of potential adverse effects of public health interventions developed by Lorenc and Oliver [3], and the process evaluation associated with the trial of the IHC primary school intervention [6].

We developed the criteria for a sensible framework to inform our judgements about what to include in the framework and how, for example whether to specify a potential effect or include it implicitly. Such judgements are subjective and involve trade-offs. The criteria help make those judgements explicit, transparent, and systematic. We based the initial criteria for a sensible framework on relevant criteria for sensible health indexes developed by Feinstein [19], since in previous work, we and others have found them to be a helpful starting point for developing various tools [20, 21].

### Survey of experts, and revision of the criteria and framework

We developed a survey (S2 File) for collecting expert feedback on the first version of the framework and criteria for a sensible framework, using Nettskjema, an online survey tool developed and hosted by the University of Oslo [22]. The survey included both Likert-scale items, for collecting broad, quantitative data, and open-ended questions, for collecting more nuanced, qualitative data.

MO emailed a request for feedback to 70 external experts, as well as the research team. He attached the initial framework and criteria (S1 File) and included a link to the survey (S2 File). The external experts included health and education researchers; other non-researcher educationalists, including teachers and curriculum developers; and information researchers and designers, based in low-, middle-, and high-income countries. They included all 51 members of our international advisory network, and 19 other external experts with particularly relevant expertise. The latter were recommended by members of the research team or advisory network, or early respondents to the survey.

MO conducted a thematic analysis of the qualitative survey data [23]. The analysis included five steps:

1. Reviewing all the qualitative data and tagging each data point with an initial theme, in a spreadsheet
2. Reviewing and revising the tags and themes
3. Organising the data by theme, in a document
4. For each theme, suggesting what changes to make to the criteria or framework, if any
5. Revising the analysis based on feedback from co-investigators

MO then made the agreed-upon revisions to the criteria and framework.

### Interviews with teachers, and prioritisation of outcomes

We conducted individual interviews with teachers to identify any potential adverse effects missing from the revised framework and preliminarily prioritise potential adverse outcomes in the framework to explore in the process evaluations and measure in the one-year-follow-up assessments of the trials (Box 1) (Fig 1). We recruited one teacher each from Rwanda, Kenya, and Uganda.

We included teachers familiar with the project through participation in the IHC teacher network in their country [24] and piloting prototypes of the digital resources [11]. We excluded teachers unfamiliar with the project since they were unlikely to have experience teaching critical thinking based on findings from the context analyses (Box 1) [12–14], consistent with findings from similar studies in Norway and Australia [25–27].

We developed an interview guide that we revised after each interview. S3 File is the final version of the guide. We expected the interview topic—potential adverse effects (“disadvantages”) of teaching critical thinking—would be unfamiliar or strange to teachers, given that adverse effects of educational interventions are often overlooked [1]. Moreover, we expected that they might find the topic uncomfortable, since they might experience it as being asked to criticise themselves, their colleagues, or education authorities. Therefore, we first asked about potential beneficial effects (“advantages”) of teaching critical thinking. Furthermore, we opted for individual interviews, rather than group interviews, since individual interviews are logically more appropriate for exploring knowledge that is taken for granted and not readily articulated [28].

MO led the interviews via video chat, using Zoom. In two of the three interviews, at least one of the IHC team members in the relevant country joined in-person (RS in Uganda) or via video chat (FC and BN in Kenya), to observe and take additional notes. We video-recorded all the interviews.

MO entered all notes into a spreadsheet, adding transcriptions of the participants’ responses to questions about potential adverse effects of teaching critical thinking. We linked those responses (data points) to relevant outcomes in the framework. If the teacher reported experiencing or observing a specific adverse outcome during a pilot of the prototype resources or—explicitly or implicitly—suggested a specific adverse effect of the intervention was possible, we preliminarily prioritised the outcome for evaluation.

### Ethical considerations

Participation in this study was voluntary and did not involve likely or serious risks to participants.

Before publishing the article, we sent the manuscript to the survey respondents who provided feedback on the initial framework. We asked them to contact us if they had any concerns. We have not published data that could be used to individually identify them, and we have removed the names of colleagues mentioned in their responses. We did not obtain consent from the survey respondents because the data were analysed anonymously. We obtained written consent from the interview participants for the entire development stage of the project, including being recorded.

The Norwegian Institute of health (NIPH) is the project’s lead partner. As required by NIPH— to comply with the European General Data Protection Regulation—we have completed a data privacy impact assessment (DPIA) for the entire development stage of the project, including this study. The Data Protection and Chief Information Security Officers at the Norwegian Institute of Public Health (NIPH) provided feedback on the DPIA, and the relevant senior advisor at NIPH approved it. Furthermore, as required by RCN, we have created a data management plan for the entire project, which we are updating continuously and will submit to RCN at the end of the project. Since the project will not generate new knowledge about health and disease, it falls outside the remit of the Regional Committee for Medical Research Ethics [29], in Norway, which the committee has confirmed (reference number 30713).

In Kenya, we obtained ethics approval from Masinde Muliro University of Science and Technology Institutional Ethics Review Committee and the Kenya National Commission of Science and Technology Institute (licence number NACOSTI/P/19/1986), as well as approval from the Ministry of Education and the Teachers Service Commission, nationally and at the county-level. In Rwanda, we obtained ethics approval from the Rwandan National Ethics Committee. In Uganda, we obtained ethics approval from the School of Medicine research ethics committee at the Makerere University College of Health Sciences, and from the Uganda National Council for Science and Technology.

## Results

The results of this study include the following, in order: findings from the survey of experts; the revised criteria for a sensible framework; the revised framework, including an overview (a series of six tables) and descriptions of the potential mechanisms; findings from the interviews with teachers; and the prioritised outcomes.

### Survey of experts, and revision of the criteria and framework

We received 42 survey responses from experts in:

- Developing and evaluating educational interventions, in general and specifically within critical thinking
- Developing and evaluating health interventions, including public health interventions
- Teaching critical thinking, in general and specifically about health

Four of the survey respondents (10%) were members of our team. In other words, just over half of the external recipients responded to the survey (38/70, 54%). All but one of those respondents were members of our advisory network. A few of the experts we contacted chose to send qualitative feedback via email, in addition to the survey (3) or instead (4). The four who provided feedback via email instead of the survey were external and not members of the advisory network. Five of the survey respondents also provided feedback in copies of the circulated document (S1 File).

Most of the survey respondents were researchers (33 of 42, 79%). Many of those respondents were also practitioners, i.e., educators or health professionals. Six were non-researcher educationalists, including teachers and curriculum developers. In total, twenty-four (57%) of the respondents worked primarily in health—including public health, health literacy, and evidence-based health care—and 15 (36%) in education. The remaining three respondents were primarily information researchers and designers.

Thirty-one (74%) of the survey respondents were based in high-income countries, 7 (17%) in middle-income countries, and 4 (10%) in low-income countries. Twenty-four (57%) were based in Europe, 8 (19%) in Africa, 5 (12%) in North America, 2 (5%) in Australia, 2 (5%) in Central- or South-America, and 1 (2%) in Asia.

Responses to the Likert-scale items suggested respondents overall approved of the initial framework and criteria (S4 File). However, based on the analysis of the qualitative survey data, we substantially revised both. S5 File is the qualitative data organised by theme, together with our responses, including the specific revisions we made. Broadly speaking, those changes included:

- Adjusting the objectives
- Adjusting, removing, and adding criteria for a sensible framework
- Dividing the overview of the framework into a series of tables
- Adjusting outcome categories
- Adjusting or removing outcomes
- Adjusting the potential mechanisms
- Replacing problematic terms and adding definitions

### Revised criteria for a sensible framework

Box 2 shows the revised criteria for a sensible framework.

#### Box 2. Revised criteria for a sensible framework of potential adverse effects of the IHC secondary school intervention.

- Each category of potential adverse effects is clear and logical.
- Each potential adverse effect is clear, logical, and likely to be important to participants or other stakeholders.
- Each corresponding beneficial effect is clear and logical.
- The amount of content in the framework is manageable.
- The organisation of the framework is clear and logical.
- The presentation of the framework is clear and logical.

### Revised framework

The revised framework has two parts. The first part is an overview: a series of six tables presenting the categories of adverse outcomes with definitions (Table 1), outcomes within those categories (Table 2), definitions of those outcomes (.Table 3), sub-outcomes (Table 4), potentially affected individuals, groups, and populations (Table 5), and corresponding beneficial outcomes (Table 6). The second part of the framework is descriptions of the potential mechanisms for the effects, with examples, which follow the overview.

**Table 1.**
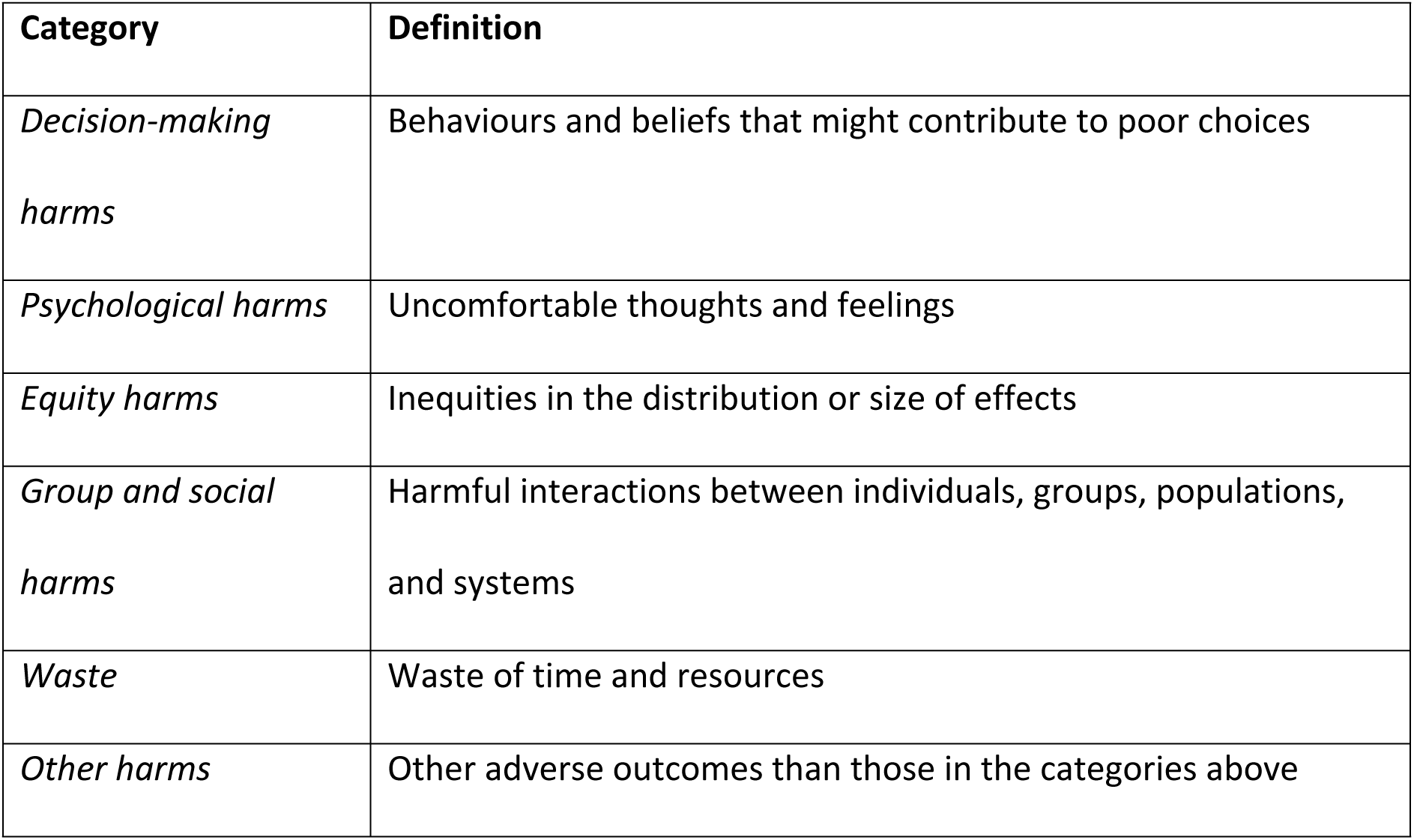
Categories of adverse outcomes.

**Table 2.** Adverse outcomes by category.

| Category | Adverse outcome |
| --- | --- |
| *Decision-making harms* | Misunderstanding |
|  | Misapplication of learning |
| *Psychological harms* | Distrust<sup>1</sup> |
|  | Pessimism<sup>2</sup> |
|  | Cognitive dissonance |
|  | Work/Schoolwork-related stress |
| *Equity harms* | Benefit-based inequity |
|  | Harm-based inequity |
| *Group and social harms* | Conflict |
| *Waste* | Wasted time or resources |
| *Any category* | Other harms than those above |

<sup>1</sup>Distrust might also be a decision-making harm, or group or social harm.
<sup>2</sup>Pessimism might also be a decision-making harm.

**Table 3.**
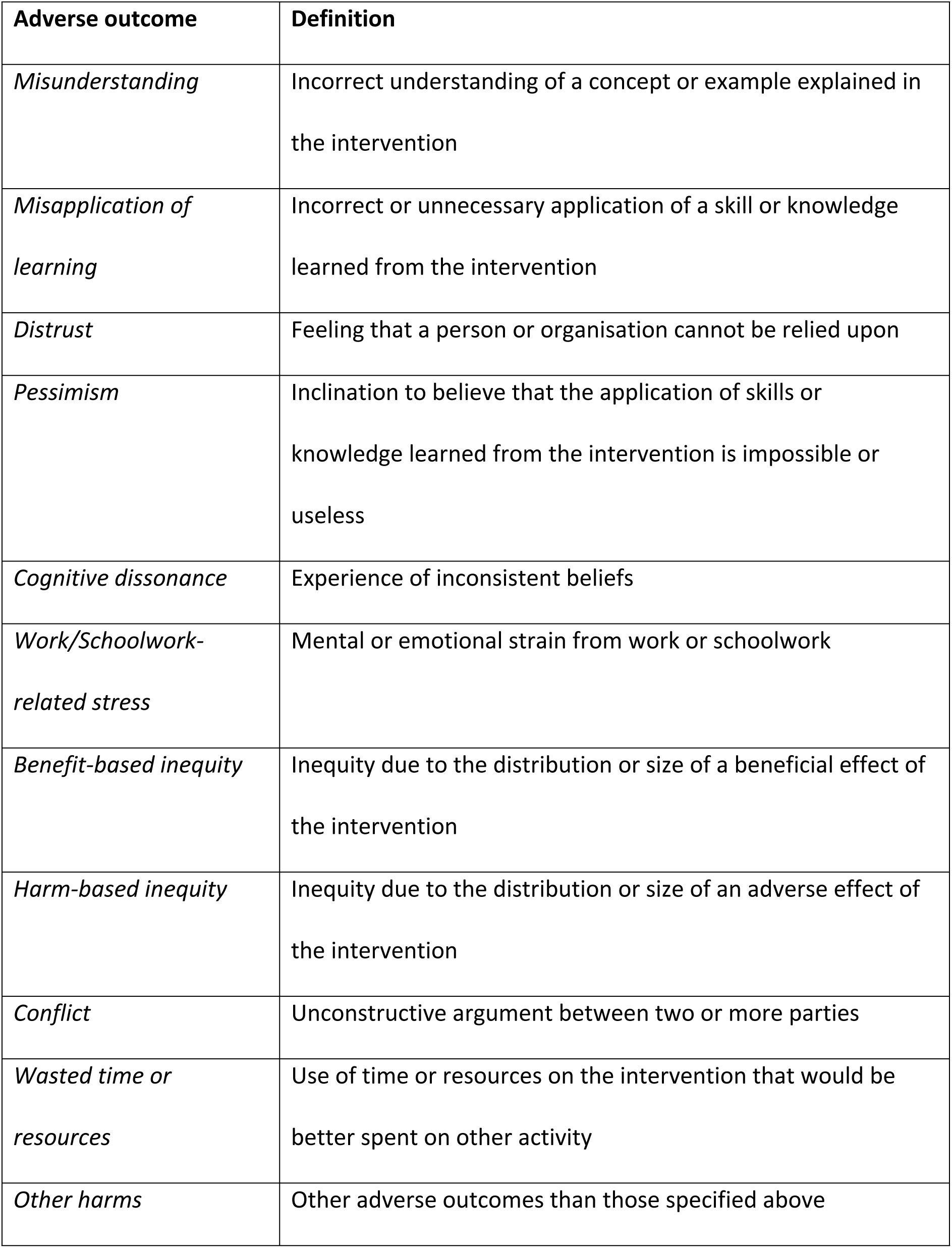
Definitions of adverse outcomes.

| **Adverse outcome** | **Definition** |
| --- | --- |
| *Misunderstanding* | Incorrect understanding of a concept or example explained in the intervention |
| *Misapplication of learning* | Incorrect or unnecessary application of a skill or knowledge learned from the intervention |
| *Distrust* | Feeling that a person or organisation cannot be relied upon |
| *Pessimism* | Inclination to believe that the application of skills or knowledge learned from the intervention is impossible or useless |
| *Cognitive dissonance* | Experience of inconsistent beliefs |
| *Work/Schoolwork-related stress* | Mental or emotional strain from work or schoolwork |
| *Benefit-based inequity* | Inequity due to the distribution or size of a beneficial effect of the intervention |
| *Harm-based inequity* | Inequity due to the distribution or size of an adverse effect of the intervention |
| *Conflict* | Unconstructive argument between two or more parties |
| *Wasted time or resources* | Use of time or resources on the intervention that would be better spent on other activity |
| *Other harms* | Other adverse outcomes than those specified above |

**Table 4.** Sub-outcomes.

| Adverse outcome | Sub-outcomes |  |
| --- | --- | --- |
| *Distrust* | towards | • Health professional<br>• Health researcher<br>• Family<br>• Teacher<br>• Other |
| *Conflict* | involving | • Participant<sup>1</sup><br>• School<br>• Family<br>• Social networks<sup>2</sup><br>• Health services<sup>3</sup><br>• Researcher<br>• Other |
| *Benefit-based inequity* | between | • High and low-achieving students<br>• High and low-resourced students<br>• High and low-resourced schools<br>• Other |

<sup>1</sup>Student or teacher
<sup>2</sup>Friends, peers, colleagues, religious leaders, and others
<sup>3</sup>Health professionals and others

**Table 5.**
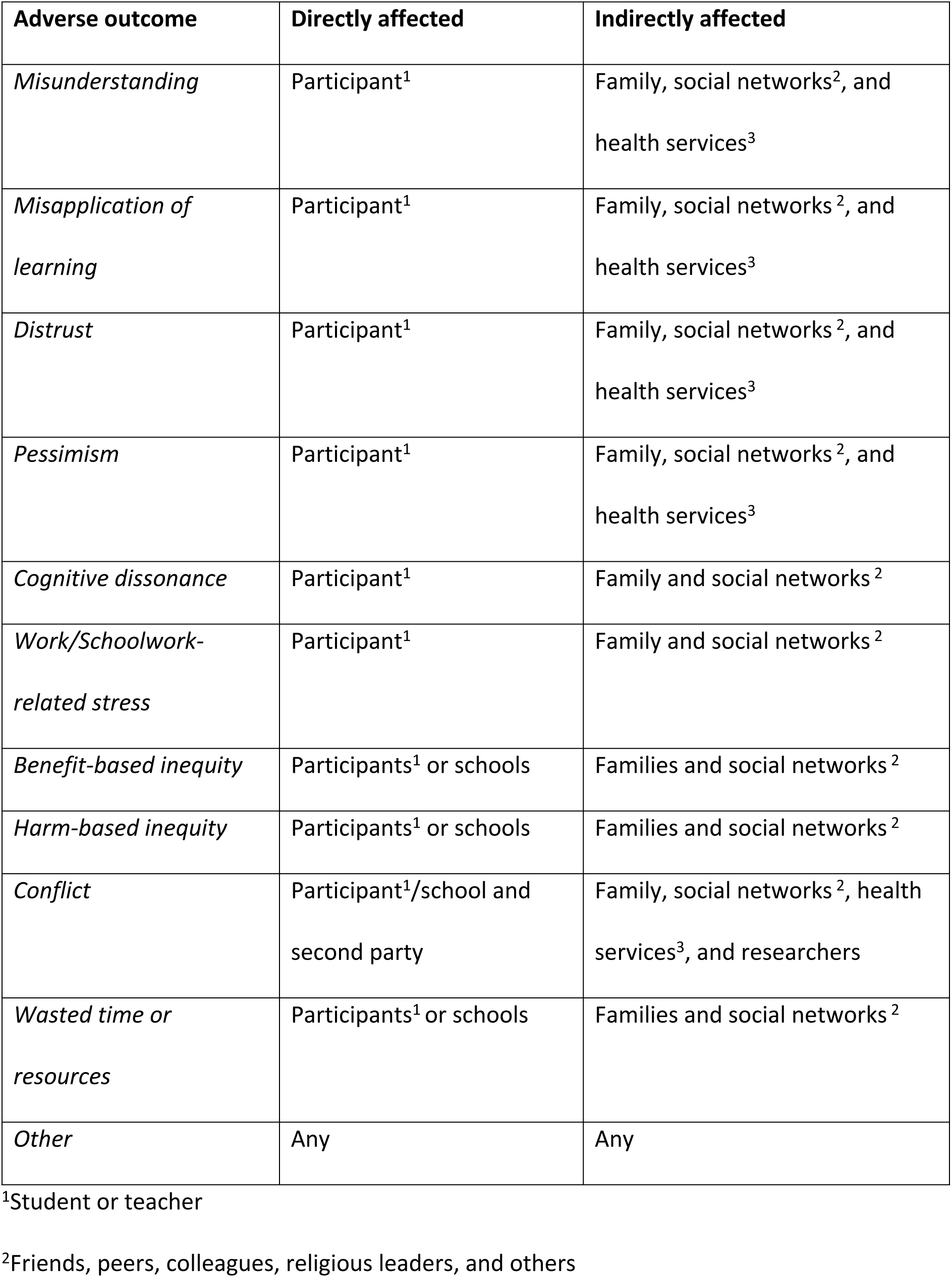
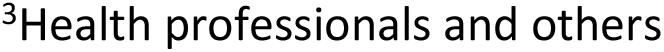
Affected individuals, groups, and populations, by adverse outcome.

| **Adverse outcome** | **Directly affected** | **Indirectly affected** |
| --- | --- | --- |
| *Misunderstanding* | Participant<sup>1</sup> | Family, social networks<sup>2</sup>, and health services<sup>3</sup> |
| *Misapplication of learning* | Participant<sup>1</sup> | Family, social networks<sup>2</sup>, and health services<sup>3</sup> |
| *Distrust* | Participant<sup>1</sup> | Family, social networks<sup>2</sup>, and health services<sup>3</sup> |
| *Pessimism* | Participant<sup>1</sup> | Family, social networks<sup>2</sup>, and health services<sup>3</sup> |
| *Cognitive dissonance* | Participant<sup>1</sup> | Family and social networks<sup>2</sup> |
| *Work/Schoolwork-related stress* | Participant<sup>1</sup> | Family and social networks<sup>2</sup> |
| *Benefit-based inequity* | Participants<sup>1</sup> or schools | Families and social networks<sup>2</sup> |
| *Harm-based inequity* | Participants<sup>1</sup> or schools | Families and social networks<sup>2</sup> |
| *Conflict* | Participant<sup>1</sup>/school and second party | Family, social networks<sup>2</sup>, health services<sup>3</sup>, and researchers |
| *Wasted time or resources* | Participants<sup>1</sup> or schools | Families and social networks<sup>2</sup> |
| *Other* | Any | Any |

<sup>1</sup>Student or teacher
<sup>2</sup>Friends, peers, colleagues, religious leaders, and others
<sup>3</sup>Health professionals and others

**Table 6.** Corresponding beneficial outcomes.

| **Adverse outcome** | **Corresponding beneficial outcome<sup>1</sup>** |
| --- | --- |
| *Misunderstanding* | Understanding |
| *Misapplication of learning* | Correct and necessary application of learning |
| *Distrust* | Healthy scepticism |
| *Pessimism* | Optimism |
| *Cognitive dissonance* | Cognitive coherence |
| *Work/Schoolwork-related stress* | None |
| *Benefit-based inequity* | Benefit-based equity |
| *Harm-based inequity* | None |
| *Conflict* | Constructive discussion |
| *Wasted time or resources* | Worthwhile use of time and resources |

<sup>1</sup>The corresponding beneficial effect (and increase in the corresponding beneficial outcome) is not necessarily the converse of the adverse effect, but a potential beneficial effect in place of the potential adverse effect.

#### Misunderstanding

The IHC secondary school intervention might cause misunderstandings of concepts or examples, which participants might then “transfer” [30] to their daily lives. Such effects are possible if the concepts or examples are unclear to participants due to a problem with how they are explained or used in the intervention (Box 3). In particular, examples of health claims might be misunderstood as advice if it is unclear how the IHC secondary school intervention is different from typical public health interventions, which encourage particular choices, such as getting screened for a disease [31], as opposed to helping people learn general decision-making skills.

##### Box 3. Examples of potential misunderstandings caused by the intervention.

**Misunderstanding a concept**

In terms of the IHC Key concepts about the reliability of claims and research evidence, a participant might misunderstand “unreliable” as meaning the same as “false” or “incorrect”. This is possible if the meaning of “unreliable” is unclear due to a problem with its explanation or use in the training or resources. In their daily life, when faced with an unreliable claim about the effects of a health intervention, the participant might then assume the claim is simply false and the intervention does not have the claimed effects. In fact, the intervention might have the claimed effects, despite the claim being unreliable. Moreover, there might be an alternate, reliable basis for the claim. For example, when faced with a claim about the helpful effects of a vaccine based on anecdotal evidence, the participant might assume the claim is false and the vaccine is wasteful, if not harmful. However, the vaccine might still be helpful, and there might be reliable evidence showing it is helpful.

**Misunderstanding an example**

It is possible that participants will interpret examples of reliable claims as advice. This is possible if the explanation or use of an example is unclear. For example, a participant might interpret an example about evidence showing helpful effects of painkillers as advice that they should always use painkillers when in pain.

#### Misapplication of learning

The intervention might improve skills in the learning context (i.e. the training or lessons), but cause misapplication of those skills in other contexts, most importantly participants’ daily lives. Such “mis-transfer” of learning—as opposed to transfer of mislearning (see “Misunderstanding”)—is possible if limitations of the intervention are not addressed clearly, in the intervention. Those limitations include that the IHC secondary school intervention focuses on a limited number of IHC Key Concepts (.Box 1), and that it does not focus on other relevant and important skill sets, such as the ability to search for relevant and reliable information.

For example, in the learning context, a participant might understand the concept that randomisation is the only way to control for unknown confounding. However, the intervention does not focus concepts about blinding. If this limitation of the intervention is unclear, when in daily life the participant is faced with evidence from a randomised, but unblind trial, with a high risk of bias, they might then assume the evidence is more reliable than it really is.

Misapplication of learning here includes “overtransfer” [30]: technically correct, but unnecessary application of learning. For example, a participant might spend time and energy assessing the basis for a health claim that could simply be ignored because the intervention in the claim is unavailable.

#### Distrust

The IHC secondary school intervention might cause participants to become distrustful of certain groups or individuals. Such effects are possible if the difference between the basis for a claim and its source is unclear to participants, as well as why the basis, not the source, determines the reliability of the claim.

For example, the intervention might cause distrust towards health professionals or researchers. This is because when explaining ways in which research can be unreliable, such as random error, we disproportionately use examples of unreliable research evaluating the effects of “Western”, “modern”, or “academic” medicine, as opposed to “traditional” or “herbal” medicine. This research typically suggests the intervention is helpful, when really it is or might be ineffective or harmful.

There are two reasons for this disproportion. First, participants might be deeply invested in beliefs about traditional and herbal care. If so, they might become defensive if we us an example suggesting such beliefs are unreliable, and their defensiveness might prevent learning. Moreover, they might assume the intervention is on the side of “Western” medicine and the pharmaceutical industry of which they might already be distrustful for understandable reasons, starting with colonialism [32].

Second, to effectively explain concepts, our findings and experiences from the development of the IHC primary and secondary school interventions suggest it is important to use real, familiar, and relevant examples, showing that the concepts are important to participants’ daily lives. However, there often is no research evaluating the effects of traditional and herbal interventions that are common in the participants’ contexts.

Increased distrust might lead to poor decisions, by causing participants to dismiss reliable and relevant advice or evidence, making it a potential decision-making harm as well as a potential psychological harm. Moreover, it might lead to conflict, making it a potential group or social harm.

In some cases, distrust might be a decision-making benefit. However, any source might provide reliable or unreliable information about the effects of a health intervention. Furthermore, aiming to increase trust or distrust in particular sources might backfire. For example, aiming to increase trust in health authorities or decrease trust in practitioners of “traditional” medicine could make the IHC secondary school intervention seem like propaganda for “Western” medicine.

Therefore, the IHC secondary school intervention is not intended to increase or decrease trust in information about particular types of interventions, nor information from particular sources. Rather, it is intended to increase “healthy” scepticism towards all claims and research evidence. If the intervention is effective, students should be more likely to seek advice from health authorities, assuming those authorities provide reliable information. And they should be less likely to seek information from sources that more often provide unreliable information.

#### Pessimism

The IHC secondary school intervention might cause participants to feel pessimistic. Such effects are possible if it is unclear to participants how the intervention can help them make better choices in many cases, even though there are relevant and important problems that the intervention cannot solve. Those problems include:

- Many common health interventions are harmful or wasteful, or have highly uncertain effects [33, 34].
- Many health claims are unreliable [35].
- Many people are limited in their ability to think critically about health information and choices, including health professionals [36–40].
- Many health research studies are unreliable or wasteful [41, 42].
- Citizens have limited influence over decisions about health policies that affect them [43].
- Free sources of reliable health information in plain language are limited in number and quality [44].
- In school, outside of the intervention, participants have limited opportunities to learn about how to think critically about health [12–14].

We address some of these problems explicitly in the intervention, and participants might recognise others. They might believe that because of these problems, applying the skills they have learned from the intervention is impossible or useless. This might be more likely in students than teachers, given that students have less experience making independent choices.

Increased pessimism might lead to poor decisions, by causing participants to mistakenly assume an informed choice is impossible or useless, making it a potential decision-making harm as well.

#### Cognitive dissonance

The IHC secondary school intervention might lead to uncomfortable cognitive dissonance. This effect is possible if the intervention causes participants to have new beliefs that are inconsistent with prior, deep-rooted beliefs, such as beliefs about the effects of “traditional” medicine (see “Distrust”). In other words, the intervention might help participants apply a concept to some beliefs about the effects of health interventions, but not others, depending on the participant’s level of emotional, social, or cultural investment in the belief. Recognising this inconsistency might be uncomfortable.

#### Work/Schoolwork-related stress

The IHC secondary school intervention might cause work-related stress in participating teachers, and schoolwork-related stress in participating students. Such effects are possible if the intervention is ineffective, inefficient, or inessential, or experienced as such, or if it is too demanding. In the process evaluation for the trial of the IHC primary school intervention, some teachers reported an increase in stress due to the added workload, and teaching something new [6].

It might be time-consuming for participants to familiarise themselves with the content of the intervention—including terminology, concepts, and teaching and learning strategies— and the design and functionality of the digital resources. Meanwhile, there is substantial pressure on students, teachers, and schools to prepare as much as possible for official exams, which do not test the ability to apply IHC Key Concepts [12–14].

Moreover, the intervention is intended to increase students’ questioning of claims, evidence, and choices. If students more often question claims or evidence from teachers, this might lead to teachers feeling stressed.

#### Inequity

The IHC secondary school intervention might cause or increase inequity. Such effects are possible if there is an unequal size or distribution of a beneficial or harmful effect across subgroups. In particular, the intervention might increase inequity amongst students depending on their baseline academic ability, or their socioeconomic background and resources. For example, some language might be suitable for some students, but too advanced for others, or some students might receive academic support at home that others do not.

Similarly, the intervention might increase inequity amongst schools depending on available resources. For example, the intervention might have a larger beneficial effect on learning in participants at highly resourced schools with projectors or printers available. Or it might cause more work-related stress in teachers at low-resource schools who have relatively little training or support.

#### Conflict

The IHC secondary school intervention might cause conflicts between different individuals and groups. Such effects are possible if the intervention causes participants to question other people’s claims, beliefs, or choices, and those people become irritated or defensive.

In particular, the intervention might cause conflict between students and authorities, such as teachers or parents. For example, a parent might be deeply invested in beliefs about the effects of “traditional” care (see “Distrust”). If the child questions any of those beliefs— especially in a way that is experienced as disrespectful—this might lead to conflict.

#### Wasted time or resources

With any intervention, there is an opportunity cost. The IHC secondary school intervention might cause participants and schools to waste time and resources. Such effects are possible if the intervention is ineffective or inessential, or experienced as such.

As mentioned under “Work/Schoolwork-related stress”, there is substantial pressure on students, teachers, and schools to prepare as much as possible for official exams [12–14]. This is especially important in terms of waste as well, since any time spent on an ineffective or inessential intervention could have been spent on said preparation for exams.

#### Other potential adverse outcomes

The IHC secondary school intervention might have adverse effects that are not specified in the framework. This includes effects that we have considered and chosen not to specify, because they seem illogical, such as an increase in bullying (S5 File), as well as any effects that we have failed to consider.

### Interviews with teachers, and prioritisation

We interviewed one teacher in each of the three countries where we are initially evaluating the IHC secondary school intervention (Table 7). We organised the interview data in five main themes (S6 File). Within the theme about potential adverse effects (“Disadvantages”), which was the focus of the analysis, we organised the data into 11 sub-themes. Each sub-theme is a potential adverse outcome, or a factor in the potential mechanism of an adverse effect (S6 File).

**Table 7.** Interview participants.

|  | Teacher 1 | Teacher 2 | Teacher 3 |
| --- | --- | --- | --- |
| *Country* | Uganda | Kenya | Rwanda |
| *Years of teaching experience* | 10 | 4 | 10 |
| *Subjects* | Biology, chemistry | Biology, agriculture | Physics |
| *Forms (grades or years)* | 1-4 (entire lower secondary) | 1-4 (entire lower secondary) | 4-6 (entire upper secondary) |
| *School location* | Urban | Semi-urban | Urban |
| *School type* | Private | Public | Semi-public<br>(“government-aided”) |

If a teacher reported observing the outcome in the pilot, or suggested the effect is possible in the trial, we marked the outcome as a priority for evaluation, preliminarily. In the end, this led us to prioritising three outcomes: work-related stress; wasted time or resources; and conflict, particularly conflict between students and family. None of the teachers reported or suggested any potential adverse outcomes or effects that we had not already included in the framework.

Teacher 1 reported two “worries” or “challenges” for teaching critical thinking, which we linked to the outcomes “Wasted time or resources” and “Work-related stress”. Teacher 2 reported experiencing stress and wasted time in the pilot, and suggested conflict was another potential adverse effect was possible, although he had not experienced or observed it. Teacher 3 initially said there were no potential adverse effects of teaching critical thinking: *“When you think critically, you can go far, but when you don’t think critically, I think you can go nowhere. So, [there is] no disadvantage.“* When we asked him about the potential adverse effects suggested by Teachers 1 and 2, he suggested those effects were not possible, or—if possible—acceptable or easily addressed.

#### Conflict

Teacher 1 did not raise conflict as an adverse outcome potentially caused by the intervention. Although he had not observed or experienced it in the pilot, Teacher 2 suggested the intervention could cause it, specifically between students and family.

> *“These parents may be illiterate. They never went to school. And then the students maybe, they are… There is something that they do out of their personal experience in their family. And then you realise that if we now tell the students about health actions and health choices, and then now they stop doing what they used to do in their family, this maybe sometimes might start bringing wrangles between the parent and the family, and the students, because of maybe now, they are educated and they know what is to be done.”*

Teacher 3 said an increase in “debate” was possible, but not conflict.

> *“I think [conflict] is not […] a disadvantage. It’s again considered a debate. It’s a debate, and… and a debate can end up with some decisions. I think… Among those decisions, it’s not developing hatred or any… any kind of that. So, for me, it’s not a disadvantage. It’s a debate. In a debate, you can up… end up with the… the solutions. You take the good one, but not the bad one. But it’s not disadvantage for me.”*

#### Wasted time or resources

Teacher 1 expressed concern that students might be unable to transfer critical thinking skills to other contexts, specifically from lessons to daily life, and from health care to other fields. If transfer of learning from the intervention is limited, the value of the intervention is limited. Therefore, we linked this finding to “Wasted time or resources”.

> *“In fact, I have a question in mind: can this be taught? Can it be taught, teaching someone how to think critically? Yes, you put up so many scenarios [in the lessons] where one would think critically, and maybe the solution is there, but there are different situations [in daily life].”*

> *“[Because] we are saying critical thinking may not only be about health […] Critical thinking is going to help someone solve a problem in general. […] But again, you find that this will again involve someone’s ability to what? Understand these are things that are variable.”*

Her second “worry” was that critical thinking might be down-prioritised as long as it is not on an official exam. We linked this finding to “Wasted time or resources” too, since the intervention might be ineffective or inessential, in which case time spent on the intervention might be better spent preparing for such exams.

> *“You find that the respect it is given, it is not the same respect that is given to something that is to be examinated [sic] at the end of the day. And this starts from the administration. [Because] you find that in Uganda today, the curriculum—whether or old or new—that we are having… You find that we have a challenge that people are exam-oriented. Anything that is not to do with them having academic excellency would be wasting time. You realise that some schools don’t even do their sports, the what, and what, because they think that it would waste time, since at the end of the day, we don’t have an exam on sports, we don’t have an exam on [Music, Dance and Drama], we don’t have… You know?”*

Teacher 2 reported experiencing work-related stress during the pilot due to students questioning claims and becoming distracted, as well as the teacher feeling unprepared for questions from students about “traditional claims” and “taboos”.

> *“Yeah, [the students] became so inquisitive, and they also [started] ’doing their own research’ […] So this maybe take [sic] time, and again sometimes they also divert the attention of the others. Instead of them… all of them concentrating [on the same thing], they have divided attention, because somebody maybe brought something that was so interesting, out of his own critical thinking.”*

> *“So, they brought in traditional culture and some traditions, and even taboos, as claims. So, if they ask for you… ask you to clarify the claim, maybe sometimes you don’t even know… You are… You are hearing about it for the first time. So, it became challenging for us.*

> *Because some of the… the… the taboos that were said by so many old people, long, long time ago, [the students] bring them today as claims about health actions, and it was challenging for us even to answer such questions or respond to such questions.”*

When asked for an example, he said students had asked whether visiting a witchdoctor was a “health action”, which is the term used as a plain-language alternative to “health intervention” in the digital resources, defined as “something that someone does to care for their health or the health of others.” The teacher said he did not want to say that visiting a witchdoctor is a “health action”—even though it is by the definition in the resources— because he feared students would interpret it as him saying that visiting a witchdoctor is good for your health.

> *“There was a question like… We can talk about visiting a witchdoctor; is it a health claim or an [sic] health action? So, we were in between there. ’Can we talk about… Is it a health action or a health claim?’ […] We were stuck, whether to… Because visiting witchdoctors for us [teachers], it is wrong. And we did not… We cannot… We cannot say that that is an [sic] health action. […] We just said it was a claim […] [The students were] not satisfied, but we had to move on.”*

Teacher 3 agreed that the pilot lessons were “time-consuming”, but worthwhile, assuming they are effective.

> *“Teaching these… these lessons or critical thinking, it’s time-consuming. But when you spend time to teach, think critically, and then you come up with the good result, whatever the time it will take, I think it’s good, even if it’s time-consuming.”*

#### Work-related stress

In addition to “Waste of time or resources”, we linked the first concern expressed by Teacher 1—that students might be unable to transfer critical thinking skills to other contexts—to “Work-related stress”, since it might be stressful for teachers to balance the importance of teaching students to think critically with pressure to spend as much time as possible preparing for official exams. Similarly, we also linked the experience described by Teacher 2—students questioning claims and becoming distracted, and him feeling unprepared for questions from students about “traditional claims” and “taboos”—to “Work-related stress”.

Teacher 3 agreed that work-related stress was a possible effect of teaching critical thinking, because teaching critical thinking is “time-consuming” (see interview findings regarding “Wasted time or resources”) and because it might cause students to ask difficult questions to which teachers do not have the answer. However, he said teaching critical thinking was worth the time, and that a teacher could search for answers to students’ questions, possibly together with students, or consult an expert.

> *“Yeah, it’s possible. It’s possible. But when the… When the student, or a learner, ask… the… the question, or brings a claim that you don’t know… So, it’s better to do a research [sic], or even consult the expert. […] Or we can do the research together, can come up with a solution.”*

## Discussion

### Strengths

As far as we are aware, the framework presented in this article is the first tool of its kind in education research. It is informed by a substantial amount of quantitative and qualitative feedback from researchers and other experts, and interviews with teachers who have rare experience teaching critical thinking in their contexts, as well as evidence and theory from previous studies, and other tools. Moreover, it is the result of interdisciplinary collaboration, which is important when trying to help people learn how to think critically [45–47]. By developing the framework, we identified potential adverse effects that are logical and likely to be important, if they occur. Our preliminary prioritisation of outcomes is consistent with findings from the process evaluation for the trial of the IHC primary school intervention [6].

The framework highlights that there might be other adverse effects of the IHC secondary school intervention and other interventions intended to improve critical thinking, which we either considered and chose not to specify in the framework or failed to consider (see “Other potential adverse outcomes”). Moreover, the framework is a “living” tool, which can be improved upon, as well as adapted.

The framework is primarily a tool for researchers. However, it can be used as the basis for developing tools for other groups, for example evaluation forms for teachers. This would be similar to how the IHC Key Concepts framework has been used as the basis for developing the IHC primary and secondary school interventions [7,11,48,49].

### Limitations

Important limitations of the framework include limits to its comprehensiveness, and the use of terminology with different meanings or interpretations depending on the context. A limitation of this study is the limited number of interviews. Also, the generic value of the framework is uncertain, granted developing a generic framework was not our objective.

In the evaluation of the IHC secondary school intervention, we will address the limitations of the framework and this study by using complementary mixed methods (S7 File), being transparent, and showing appropriate caution in the interpretation of results. As mentioned in the background section, the prioritisation of outcomes is preliminary, and we will consider re-prioritisation during the process evaluations.

#### Comprehensiveness

Adverse effects can be complex, and we have made trade-offs between making the framework manageable versus comprehensive. We have not presented relationships between potential mechanisms in the overview (Tables 1-6) or a model, such as the potential for distrust to lead to conflict (see “Distrust”). Further, we have limited the number of specified outcomes (Table 2) and sub-outcomes (Table 4). On the other hand, in the evaluation of the IHC secondary school intervention, we do not have enough time or resources for in-depth evaluation of every outcome that is specified, which is why we are prioritising outcomes, initially based on the interviews with teachers in this study.

#### Terminology

Some of the terminology we have used will have different meanings or interpretations in other contexts. For example, “distrust”, “pessimism”, and “cognitive dissonance” are not necessarily defined, understood, or experienced as uncomfortable—they might even be beneficial, in some cases. Similarly—as mentioned by one of the survey respondents (S5 File)—misunderstandings and misapplication of learning, if effectively addressed, might ultimately aid learning, without causing harm.

#### Interview participants

It is possible that additional interviews with teachers or other stakeholders would have changed our interpretation of the data from the three interviews we did conduct, or provided additional findings. It is also possible that the three teachers who participated in this study felt invested in the project from their previous participation, and therefore were unrepresentatively positive towards teaching critical thinking and specifically our intervention. Moreover, it is possible that data were misinterpreted due to language barriers.

#### Generic value

We specifically developed the framework to inform the development and evaluation of the IHC secondary school intervention. Based in part on the survey feedback, we believe the framework can be helpfully used to develop and evaluate other interventions intended to improve critical thinking, particularly about health, but also within other fields. A majority of the IHC Key Concepts are relevant to at least 13 other fields, besides health care [46].

However, the evidence that informed the framework is mostly from IHC studies [5,6,12–14], and limited to East-African contexts. Unfortunately, there was limited evidence about adverse effects of other educational interventions to test against the framework [1, 2]. Therefore, we cannot be certain about the generic value of the framework until it is used in other projects. However, the general approach to evaluating potential adverse effects of an educational intervention can have value to other researchers regardless.

## Conclusion

In this study, we have identified potential adverse effects of the IHC secondary school intervention, by developing a framework, and we have preliminarily prioritised outcomes in the framework for the evaluation of the intervention. Overall, survey responses from researchers and others with a variety of relevant expertise were positive to an initial version of the framework. However, responses also included critical feedback that led to substantial revision. Teachers interviewed about potential adverse effects of teaching critical thinking did not report or suggest any potential adverse outcomes or effects that were missing from the framework. Based on the interview findings, we prioritised three outcomes for the evaluation of the IHC secondary school intervention: work-related stress; wasted time or resources; and conflict, especially between students and family.

In S7 File, we describe how we have used and plan to use the framework in the development and evaluation of the IHC secondary school intervention, exemplifying how it can be used in other projects. In short, we have used it to inform the development of interview and observation guides being used in the process evaluations, and we are using it to inform the development of outcome measures for the 1-year-follow-up assessments of the trials.

Rigorous evaluations of potential adverse effects of educational and public health interventions, as well as efforts to prevent those effects, can be time and resource-intensive. However, these evaluations might come at a small cost compared to the cost of implementing harmful interventions. We hope the framework developed in this study will make more efficient the prevention and evaluation of potential adverse effects of other educational interventions.

## Data Availability

All relevant data are within the manuscript and its Supporting Information files.

## Contributions

MO planned the study, drafted the initial framework, survey, and interview guide. He collected, managed, and analysed the survey data, led the interviews, and managed and analysed the interview data. He suggested changes to the framework based on the survey and interview findings. He also drafted this article. The other authors provided feedback on at least one of the following: the initial framework, the survey, the interview guide, the analysis of survey data, the suggested revision of the framework, the final framework, the analysis of interview findings, and the prioritisation of outcomes. All co-authors provided feedback on the article.

## Acknowledgements

We are grateful to the survey respondents and interview participants for their time and important insight.

## Supporting information

S1 File. Initial framework and criteria.

S2 File. Survey.

S3 File. Interview guide.

S4 File. Quantitative survey data.

S5 File. Qualitative survey data.

S6 File. Interview data.

S7 File. Use of framework.

